# Efficacy of conventional rehabilitation, transcutaneous electrical nerve stimulation, or early mobilization to reverse acquired weakness in the intensive care unit. Randomized controlled pilot trial

**DOI:** 10.1101/2023.06.05.23290971

**Authors:** César Alejandro Bueno-Ardariz, Sabrina Gimena Cagide, Claudio Gabriel Gamarra, Darío Leonel Paz, Eliana Verónica Rotela, Ricard Aranda-Castro, Fernando Daniel Bustos, Sebastián Muller, Esteban Sebastián Settembrino, Melina Carrera, Cecilia Elena Giongo, Gonzalo Martín Nannini, Paula Gimena Nardelli, Ladislao Pablo Diaz-Ballve

## Abstract

**Objective:** To compare the efficacy of conventional rehabilitation, transcutaneous electrical nerve stimulation, and early mobilization in reducing the time needed to reverse intensive care unit-acquired weakness (ICUAW), as assessed by the Medical Research Council muscle strength scale (MRC-MSS) score, in patients clinically diagnosed with ICUAW.

**Setting:** Medical-surgical ICU of a general acute care hospital with 26 beds.

**Design:** Pilot trial with random assignment and a control group.

**Interventions:** Group 1: standard or routine rehabilitation, Group 2: transcutaneous electrical nerve stimulation, and Group 3: early mobilization protocol.

**Subjects:** Individuals over 18 years old admitted to the ICU with invasive mechanical ventilation for more than 24 hours and diagnosed with ICUAW defined as Score MRC-MSS<48 measured over two consecutive days.

**Results:** 18 subjects were included. One-way ANOVA showed differences in the treatment days needed to achieve an MRC-MSS ≥48 (p = 0.01), as well as in the number of sessions required to reverse ICUAW (p = 0.01) when comparing the different interventions. Through post hoc analysis, the group treated with transcutaneous electrical nerve stimulation was identified as requiring significantly more days and sessions to reverse ICUAW. Differences were observed in the Barthel index at the end and in the variation of the Barthel index as a result of functional capacity among the different treatments.

**Conclusion:** The group treated with transcutaneous electrical nerve stimulation required a significantly greater number of days and sessions to achieve an MRC-SS ≥48. No differences in functional limitation were observed. These findings should be corroborated in similar studies. (Clinicaltrial.gov NCT04613908)

## Introduction

Due to advances in knowledge and with the support of new technologies, the survival rate of critically ill patients in the intensive care unit (ICU) has increased. However, increased survival leads to prolonged stays in the ICU with detrimental consequences for patients, such as the development of intensive care unit acquired weakness (ICUAW). ICUAW is defined as a decrease in muscle strength, typically associated with atrophy, that has an acute, diffuse, symmetrical, and generalized onset following the onset of a critical illness without any other identifiable cause. (1)(2)(3) The development of weakness is associated with increased morbidity, functional impairment, and longer hospitalization periods. (4)(5)

The pathophysiology of ICUAW is often multifactorial, with factors such as hyperglycemia, poor nutritional status, and prolonged immobilization playing a role. Additionally, the use of medications such as corticosteroids, sedatives, and neuromuscular blockers is often associated. (6)(7)(8)

The diagnosis of ICUAW can be made using different methods, with the most commonly used in the ICU being the clinical evaluation using the Medical Research Council muscle strength scale (MRC-MSS). (9) The proposed scale is simple to administer and has great clinical potential as it has been reported to have excellent reproducibility and good agreement among evaluators. (10)(11)(12) The score ranges from 0 to 60 points, with a cutoff point of <48 points to define ICUAW. The main limitation of this method is the need for patient attention and cooperation, as it is not applicable to sedated patients, those with altered consciousness, or those unable to understand or execute simple commands.

ICUAW is a condition with a high incidence, ranging from 26% to 67% in awake patients who received mechanical ventilation for more than 7 days. Therefore, once established, it must be treated emphatically to prevent its sequelae. (13)(14) The most widely used intervention is motor physiotherapy, either in a conventional form or through early mobilization strategies. (15)(16)(17) Its benefits include increased muscle strength, reduced inflammation, improved patient mood and less fatigue, leading to improvements in exercise capacity and functional status at hospital discharge. Consequently, it reduces mechanical ventilation time and hospital stay. (15)

A particularly relevant topic related to ICUAW is prevention, for which various interventions have been tested, such as transcutaneous nerve electrostimulation (TENS). TENS involves the generation of visible muscle contractions through electrical stimuli applied via surface electrodes. TENS has been shown to induce skeletal muscle growth, as well as increased power and endurance, causing a systemic effect on microcirculation and providing structural and functional benefits in critically ill patients. (18)(19)(20)(21) Despite the abundant evidence supporting its application for preventing the development of ICUAW, it has never been used as a method to reverse this condition once established. This knowledge gap led us to conduct the present study. Our aim is to evaluate the efficacy and time required to reverse ICUAW (achieve an MRC-MSS score ≥ 48) according to the applied rehabilitation treatment.

## Materials and Methods

### Setting

Medical-surgical ICU in a general acute care hospital (26 beds).

### Design

Single-blind, single-center randomized controlled trial.

The research protocol was submitted and approved by both the ethics committee and the teaching and research committee of our institution and registered at clinicaltrial.gov (NCT: NCT:04613908). Both committees authorized the conduct of the study.

Informed consent was obtained from each included subject. Subjects: 18 patients were consecutively recruited (6 per group) with a clinical diagnosis of ICUAW (MRC-MSS <48 points on 2 consecutive measurements) admitted to the ICU during the period from August 2015 to August 2016.

### Eligibility criteria

Subjects over 18 years of age, admitted to the ICU who have received invasive mechanical ventilation for a period exceeding 24 hours, and with a clinical diagnosis of ICUAW (MRC-MSS <48 measured for two consecutive days).

Subjects with a body mass index > 35 (weight in kg/height in meters squared), presence of edema that prevents the performance of TENS at the time of ICUAW diagnosis (assessed by the absence of visible contraction), pregnant women, subjects with pacemakers, subjects with central and/or peripheral nervous system injury (with motor sequelae), pre-existing neuromuscular disease, movement limitation of any limb due to traumatic or orthopedic causes, a Barthel index score of less than 35 points on admission, and those who refused to provide informed consent were excluded.

### Predictive variables

Age, sex, number of days of invasive mechanical ventilation until the first MRC-MSS measurement, Barthel index score at baseline, MRC-MSS score at the beginning, MRC-MSS score at the end of the study, MRC-MSS variation (ΔMRC), reason for ICU admission (medical or surgical), subject’s destination upon ICU discharge (death, transfer to another ward, hospital discharge).

### Outcome variables

Number of days until reaching an MRC-MSS score ≥ 48 points.

Number of sessions until reaching an MRC-MSS score ≥ 48 points.

Number of subjects per group who achieved a score ≥ 48 points on the MRC-MSS.

Barthel index score taken at the end of the 10 treatment sessions. Variation in the Barthel index (ΔBarthel), calculated as the difference between the Barthel index score before ICU admission and the score at the end of treatment.

### Procedure

Once enrolled in the study, subjects were randomly assigned to each treatment group using a random number table. Three study groups were formed as follows:

Group 1 (GR-STD) received standard or usual rehabilitation treatment used in our ICU as the intervention.

Group 2 (GR-TENS) received transcutaneous nerve electrostimulation (TENS) sessions in addition to standard treatment.

Group 3 (GR-EM) received an early mobilization protocol as the intervention.

All groups started receiving the intervention the day after randomization. The total duration of treatments was 10 sessions, once a day for 5 days a week, excluding weekends. The intervention was discontinued only when the subject experienced defined intolerance, such as refractory hypotension and/or cardiopulmonary arrest, or in the case of GR-TENS, pain that prevented the continuation of transcutaneous electrostimulation application. Subjects who missed two consecutive sessions or those who were discharged before the end of the protocol were excluded.

The application of the different interventions was performed consecutively during the same time range, always in the afternoon shift, and conducted by the same previously trained physiotherapists.

All evaluations were performed by a blinded assessor who was unaware of the type of treatment applied to the subject, to avoid assessment bias. Daily MRC-MSS measurements were recorded on a pre-designed form. The Barthel index was assessed upon ICU admission, considering the week prior to admission and ending after the 10 sessions. The details of the interventions are described in Appendix 1.

## Measurements

### Muscle strength

Muscle strength was assessed using the Medical Research Council Scale, (9) which evaluates the strength of 6 muscle groups bilaterally, 3 upper limb groups (arm, forearm, and wrist) and 3 lower limb groups (leg, knee, and foot).

The minimum evaluation is 0 (zero) if there is no contraction, with a maximum of 5 (five) if there is active movement against gravity and full resistance.

This scale is commonly used to assess ICUAW, as it allows for categorization in a simple way, differentiating them in a clinically significant manner. It has a maximum score of 60 points and a minimum score of 0 points. A score below 48 points is used as a criterion for the clinical diagnosis of ICUAW. (22)

### Functional independence

This was assessed using the Barthel Index, considering 2 measurements (at entry and at the end of the study). A cutoff above 35 on the Barthel Index was considered for functional independence.

### Sample size and power calculation

For this study, a sample size of 18 subjects was calculated (6 subjects per intervention group). The estimation was performed for a one-way ANOVA test, assuming a minimum difference of 2 sessions between the treatments, a standard deviation of 1.8 sessions, an alpha error of 5%, and a power of 80%. These values were derived from a pilot test conducted with one subject in each intervention.

### Statistical methods

Numeric variables are presented as mean and standard deviation or median and interquartile range depending on the distribution. Categorical variables are presented as frequency and cumulative percentage. Student’s t-test for independent samples or the Mann-Whitney U test was used for the comparison of continuous variables, depending on the distribution presented. The chi-square test or Fisher’s exact test was used for categorical variables, depending on the cross-tabulation table configuration. The Shapiro-Wilk test was used to assess whether the distribution of the numeric variables fit a normal distribution. Equality of variances was evaluated using the Levene’s test.

Comparison of the time to reach an MRC-MSS ≥ 48 among the groups with different interventions was analyzed using one-way ANOVA or the Kruskal-Wallis test, as appropriate. For post hoc analysis, the Games-Howell test and Dunn’s-Bonferroni pairwise comparison were used for ANOVA or Kruskal-Wallis, respectively, when these reached statistical significance. All analyses were performed using IBM SPSS Statistics for Windows, Version 22.0 (Armonk, NY: IBM Corp). A p-value <0.05 was considered statistically significant.

## Results

A total of 18 subjects were included and randomly assigned to three different treatment groups, 6 subjects per group (Figure 1). The characteristics of the complete sample are shown in Table 1, and the reasons for initiating mechanical ventilation are detailed in Table 2.

**Table 1.**
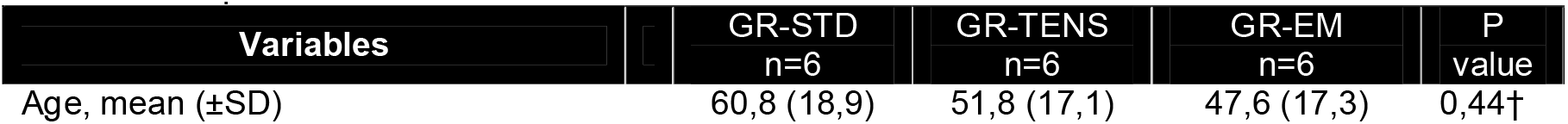
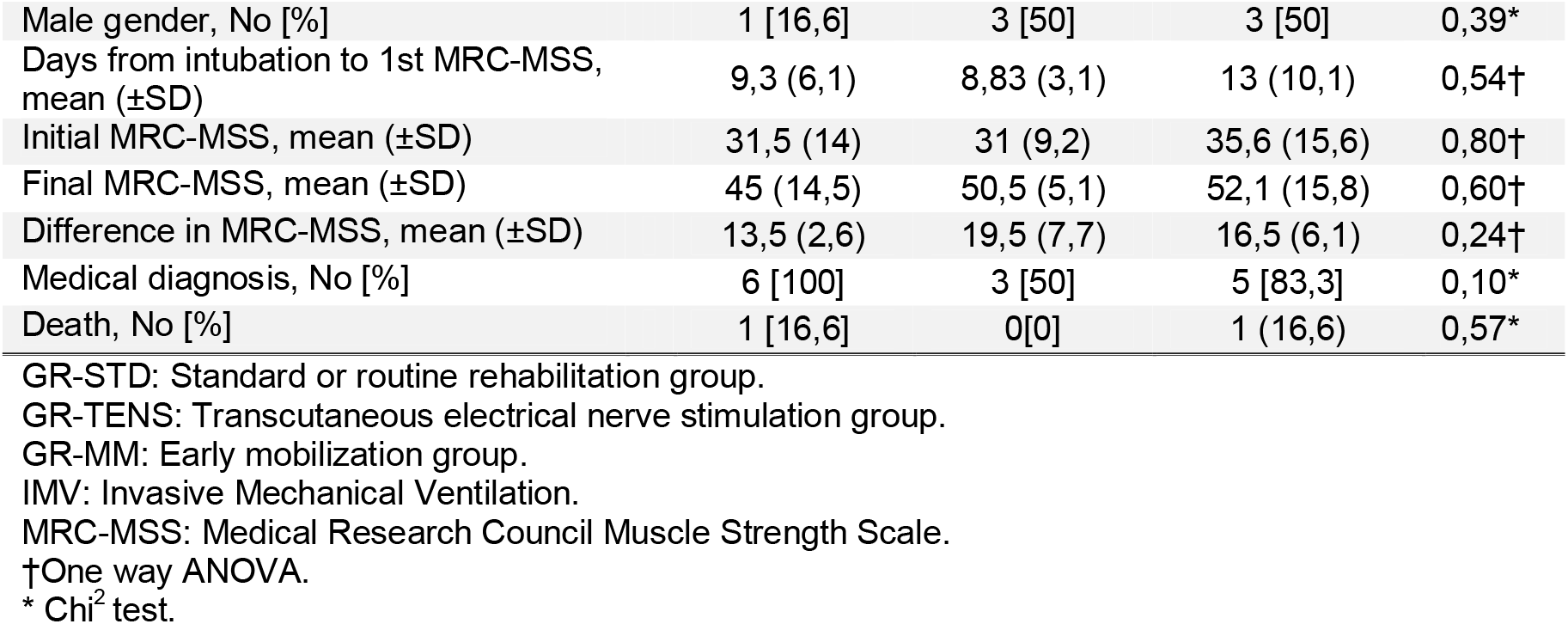
Sample Characteristics.

| Variables | GR-STD<br>n=6 | GR-TENS<br>n=6 | GR-EM<br>n=6 | P<br>value |
| --- | --- | --- | --- | --- |
| Age, mean (±SD) | 60,8 (18,9) | 51,8 (17,1) | 47,6 (17,3) | 0,44† |
| Male gender, No [%] | 1 [16,6] | 3 [50] | 3 [50] | 0,39* |
| Days from intubation to 1st MRC-MSS, mean (±SD) | 9,3 (6,1) | 8,83 (3,1) | 13 (10,1) | 0,54† |
| Initial MRC-MSS, mean (±SD) | 31,5 (14) | 31 (9,2) | 35,6 (15,6) | 0,80† |
| Final MRC-MSS, mean (±SD) | 45 (14,5) | 50,5 (5,1) | 52,1 (15,8) | 0,60† |
| Difference in MRC-MSS, mean (±SD) | 13,5 (2,6) | 19,5 (7,7) | 16,5 (6,1) | 0,24† |
| Medical diagnosis, No [%] | 6 [100] | 3 [50] | 5 [83,3] | 0,10* |
| Death, No [%] | 1 [16,6] | 0[0] | 1 (16,6) | 0,57* |

GR-STD: Standard or routine rehabilitation group.
GR-TENS: Transcutaneous electrical nerve stimulation group.
GR-MM: Early mobilization group.
IMV: Invasive Mechanical Ventilation.
MRC-MSS: Medical Research Council Muscle Strength Scale.
†One way ANOVA.
\* Chi<sup>2</sup> test.

**Table 2.** Reasons for Initiation of Invasive Mechanical Ventilation

| Reason for initiation of invasive mechanical ventilation | n | % |
| --- | --- | --- |
| Airway obstruction due to stent migration | 1 | 5.6 |
| Asthma | 2 | 11.1 |
| Chronic obstructive pulmonary disease | 3 | 16.7 |
| Acute respiratory failure |  |  |
| Community-acquired pneumonia | 2 | 11.1 |
| Status epilepticus | 1 | 5.6 |
| Nosocomial pneumonia | 2 | 11.1 |
| Massive pulmonary embolism | 1 | 5.6 |
| Postoperative | 3 | 16.7 |
| Sepsis | 4 | 22.2 |
| Total | 18 | 100 |

**Figure 1:**
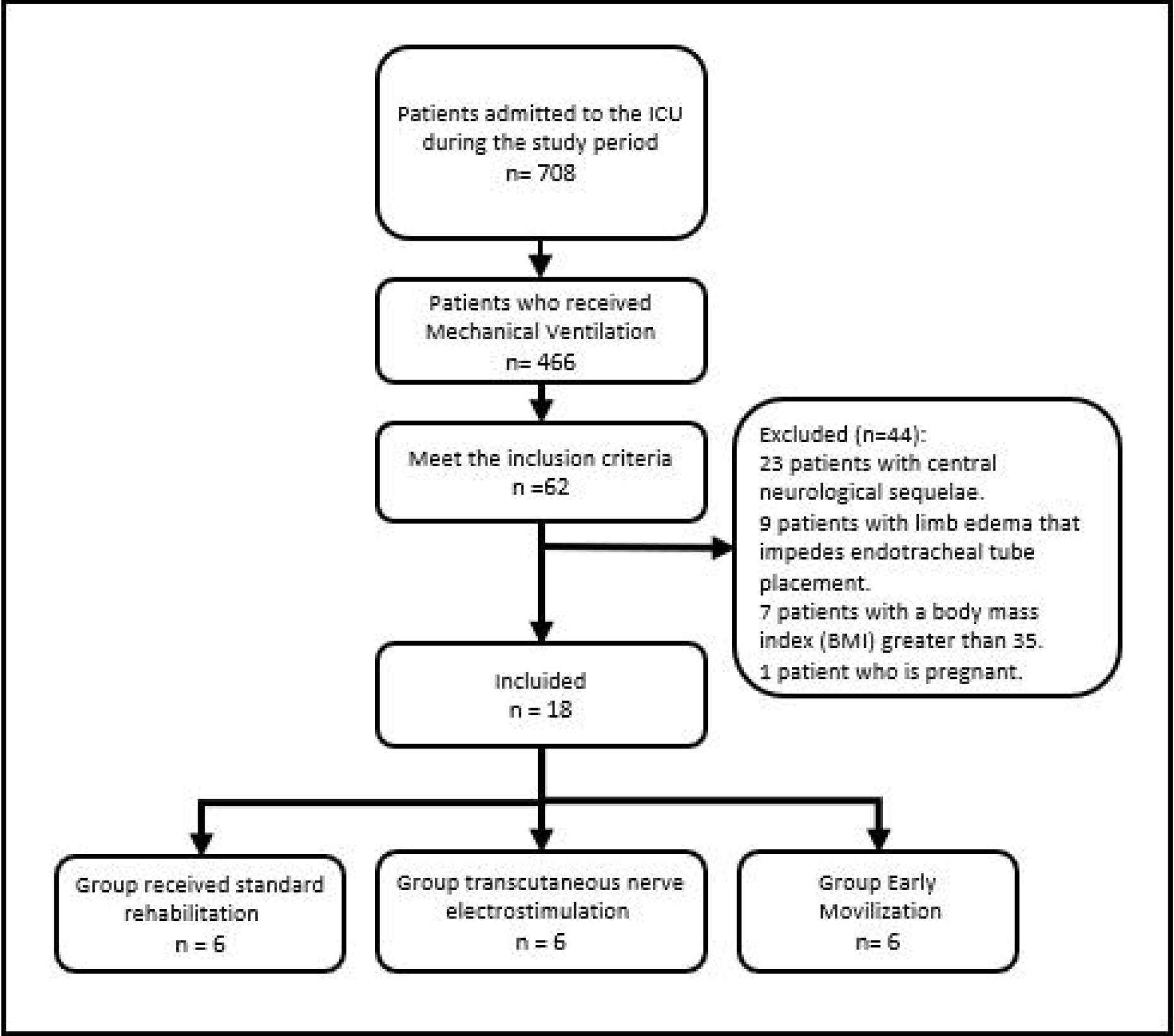
Flowchart of the patients included in the study.

At the end of the assigned treatment period (10 sessions), there were no differences in the number of subjects per group who reached an MRC-MSS ≥ 48, with four subjects in GR-STD (66.6%) and 5 in both GR-TENS and GR-EM (83.3%) (p = 0.753). The remaining subjects (2 subjects in GR-STD, 1 subject in GR-TENS, and 1 subject in GR-EM) did not reach the value of 48 in the MRC-MSS at the end of the intervention period.

Table 3 shows the one-way ANOVA comparison for the main outcome variables. Post hoc pairwise analysis for the main outcome variables that were statistically significant in the analysis of variance is presented in Table 4.

**Table 3.** Comparison of Outcome Variables by Intervention.

| Variable | GR-STD<br>n=4 | GR-TENS<br>n=5 | GR-EM<br>n=5 | P<br>value |
| --- | --- | --- | --- | --- |
| Number of days to MRC-MSS ≥48 | 4,5±2,6 | 12,8 ±2,3μ | 5,6 ±3,7 $\mu$ | 0,01 |
| Number of sessions to MRC-MSS ≥48 | 4,7 ±2,2 | 8,8 ±0,8 $\mu$ | 3,4 ±3,2 $\mu$ | 0,01 |
| Final Barthel Index | 52,5 ±35,4 | 61,6 ±38,9 | 76,6 ±39,9 | 0,74 |
| Barthel Index Difference | 47,5 [15:77,5] | 35 [3,7:73,7] | 2,5 [0:51,2] | 0,55 |

GR-STD: Standard or routine rehabilitation group
GR-TENS: Transcutaneous electrical nerve stimulation group
GR-MM: Early mobilization group
MRC-MSS: Medical Research Council Muscle Strength Scale
Note: Values are presented as mean ± standard deviation unless otherwise indicated.

**Table 4.** Post-hoc Comparison between Different Treatment Groups.

| Variable | Intervention Group |  | Mean Difference | 95% CI |  | p-value |
| --- | --- | --- | --- | --- | --- | --- |
|  |  |  |  | Inf. limit | Sup. limit |  |
| No. of days to MRC-MSS $\geq 48$ | GR-STD | GR-TENS | -8,300 | -13,95 | -2,65 | <0,01 |
|  |  | GR-EM | -1,100 | -6,75 | 4,55 | 0,92 |
|  | GR-TENS | GR-STD | 8,300 | 2,65 | 13,95 | <0,01 |
|  |  | GM | 7,200 | 1,86 | 12,54 | <0,01 |
|  | GR-EM | GR-STD | 1,100 | -4,55 | 6,75 | 0,92 |
|  |  | GR-TENS | -7,200 | -12,54 | -1,86 | <0,01 |
| No. of sessions to MRC-MSS $\geq 48$ | GR-STD | GR-TENS | -4,05 | -8,43 | 0,33 | 0,07 |
|  |  | GR-EM | 1,35 | -3,03 | 5,73 | 0,77 |
|  | GR-TENS | GR-STD | 4,05 | -0,33 | 8,43 | 0,07 |
|  |  | GR-EM | 5,4 | 1,26 | 9,54 | <0,01 |
|  | GR-EM | GR-STD | -1,35 | -5,73 | 3,03 | 0,77 |
|  |  | GR-TENS | -5,4 | -9,54 | -1,26 | <0,01 |

MRC-MSS: Medical Research Council Muscle Strength Scale.
GR-STD: Standard or routine rehabilitation group.
GR-TENS: Transcutaneous electrical nerve stimulation group.
GR-MM: Early mobilization group.
\*Gabriel's test for multiple comparisons

## Discussion

The main result found suggests that subjects treated in the GR-TENS required a greater number of treatment days to reach an MRC-MSS value ≥ 48, compared to the treatments performed in the GR-STD and the GR-EM.

In a study conducted by Routsi et al., TENS was applied and higher MRC-MSS values and fewer days of mechanical ventilation weaning were found in the treatment group. (22) In contrast, we found that subjects in the GR-TENS required a greater number of days and sessions to reach an MRC-MSS value ≥ 48. This finding suggests the limitation of TENS treatment once ICU-acquired weakness (ICUAW) has been established. However, there is evidence of improvement in exercise strength and capacity with TENS in different populations. (20) These findings were not replicated in our sample. So far, we believe that the best indication for TENS in ICU patients is in the prevention of ICUAW, which is supported by evidence such as the study by Rodríguez et al., who found increased muscle strength with daily use of TENS in sepsis patients. (18)

In the post-ICU period, we still do not have a therapeutic approach that has shown greater efficacy. Connolly et al., through a systematic review and meta-analysis, observed the effect of an exercise program in the post-ICU period on the improvement in quality of life, physical function measured by the Barthel index, muscle strength using MRC-MSS and dynamometry, as well as respiratory muscle strength. Due to the heterogeneity of the intervention and measurements, it was not possible to reach a conclusion. (21)

Schweickert et al. analyzed the survival of subjects admitted to the ICU and found that 65% of individuals who required prolonged mechanical ventilation had motor limitations at discharge. They also observed that older subjects continued to have motor limitations even one year after discharge. (23) In line with what these authors observed, we found that the Barthel index improved in 27.7% of subjects between the beginning and the end of treatment, recovering their level of functional independence at the end of the study. However, there were no differences in the Barthel index between treatments at the end of the study, suggesting that ICUAW improves over time regardless of the treatment applied. This last point is difficult to evaluate considering that the minimum standard of care for any patient with ICUAW requires some type of intervention. Based on our results, the best choice would be an early mobilization protocol or standard physiotherapy treatment. The use of TENS as a treatment to reverse ICUAW requires further studies to find benefits that justify its use in the same way as the other treatments investigated here.

Currently, several lines of research point towards bundles or packages of measures instead of isolated interventions, and it is suggested that the use of these bundles influences outcomes with high clinical relevance such as mortality, ICU days, and days of mechanical ventilation, among others. Regarding the prevention and treatment of ICUAW, bundles include early mobilization as the intervention of choice. This recommendation is based on studies that have found direct benefits with early rehabilitation (23)(24)(25) in functional outcomes at discharge. Additionally, early mobilization is the only intervention that has shown a decrease in the duration of delirium, (26) a condition that, like ICUAW, has a high incidence in ICU patients.

### Limitations

The main limitation of the study arises from the chosen evaluation method. The MRC-MSS has significant advantages that have already been mentioned, such as its applicability and cost-effectiveness, but it requires awake and cooperative patients to be implemented. As a result, the population that can be inferred is limited. There is a considerable number of patients who have ICUAW but cannot be evaluated using the MRC-MSS. It is important to point out that patients who are not able to contract a given muscle group during an evaluation are not classified as being with the muscle group compromised. In addition, the use of a single-site study can lead to selection bias in the sample studied, which may limit the generalization of the results.

## Conclusion

Based on our results, the use of TENS in the treatment of ICUAW did not show significant benefits compared to standard physiotherapy treatment or resistance exercise training. The TENS group required a greater number of treatment days and sessions to reach an MRC-MSS value ≥ 48. These findings suggest the limitation of TENS as a treatment once ICUAW has been established. However, further studies are needed to determine the benefits of TENS in the prevention of ICUAW. In the post-ICU period, there is still no therapeutic approach that has demonstrated greater efficacy, and the best choice appears to be early mobilization protocols or standard physiotherapy treatment.

## Data Availability

All data produced in the present study are available upon reasonable request to the authors

### APPENDIX 1

#### DETAILED DESCRIPTION OF INTERVENTIONS BY GROUP

Group 1 (GR-STD) received the standard or usual rehabilitation treatment used in our ICU as an intervention.

It consists of the usual treatment performed by the physiotherapist or intensive care physiotherapist. The usual strategy involves passive mobilizations in patients with functional impotence that prevents them from performing active movements (in supine position, 5 to 10 repetitions per limb in flexion, extension, abduction, and adduction).

For those patients who can perform or collaborate with active movement, their voluntary participation is requested for active/assisted movements (in supine position, 5 to 10 mobilizations per limb).

According to the professional’s judgment, assistance is provided in the seated position at the edge of the bed, and progression is based on the patient’s response, including sitting up, standing if possible, and eventually walking within the room.

This intervention was applied once a day for 2 weeks (except weekends), and unlike the early mobilization protocol, the progression criteria were the responsibility of the attending professional.

Group 2 (GR-TENS) received sessions of neuromuscular electrostimulation in addition to the standard treatment.

They received 5 sessions per week (except weekends) for two weeks using neuromuscular electrostimulation for 30 minutes each session.

The application area was prepared (shaved and cleaned with 70% alcohol-soaked cotton) and electrodes were placed. The electrode placement was similar for all patients, targeting the anterior flexor muscle groups of the shoulder and elbow (front of the arm) and the hip flexor and knee extensor (anterior thigh).

The subject was placed in a supine position with the head of the bed at 45°, legs in a neutral position, and arms at the sides of the body. We used 2 square self-adhesive electrodes of 5 cm each for the brachial biceps and 2 electrodes for each quadriceps, with the upper rectangular electrode measuring 10 cm in length and 5 cm in width, and the lower square electrode measuring 5 cm in crest.

One electrode was placed at the motor excitation point of the muscle group, and the other at the upper third of the muscle to be treated. The current used was a balanced symmetric rectangular biphasic waveform with 300-microsecond pulses and a frequency of 50 Hz, set to an intensity of up to 80 milliamperes. (26, 27, 28)

The motor excitation point was identified according to the maximum visible contraction with the lowest possible intensity. The intensity was adjusted session by session to achieve a visible contraction without generating pain or discomfort. This treatment was discontinued when visible signs of muscle fatigue, such as fasciculations, appeared.

The EET session and the standard treatment, described above, were carried out.

**(Table 5).** Dosage of NMES.

**Dosage of NMES (Table 5)**
|  |  |
| --- | --- |
| PULSE WIDTH | 300 MICROSECONDS |
| FREQUENCY | 50 HZ |
| WAVEFORM | BALANCED SYMMETRIC BIPHASIC RECTANGULAR.. |
| INTENSITY | UP TO 80 MA. |
| MUSCLES | QUADRICEPS, BRACHIAL BICEPS. |

We used a com mer cial electrostimulation device, DEMAX® Quatrum Duo model (Industria Argentina, 2014), with a pulse width of 300 microseconds, a frequency of 50 Hz, a balanced symmetric biphasic rectangular waveform, and an intensity of up to 80 mA. The intensity was adjusted session by session to achieve a visible contraction without generating pain or discomfort. This treatment was discontinued when visible signs of muscle fatigue, such as fasciculations, appeared.

Group 3 (GR-EM) received an early mobilization protocol as an intervention.

The protocol, based on the study by Morris et al (1), consisted of applying a specific treatment based on different levels of intervention for each subject. Admission to the levels was determined based on each subject’s maximum capacity, with the conditions for each level as follows:

- Level 1: MRC (shoulder) <3 + MRC <3 (hip).
- Level 2: MRC (shoulder) ≥ 3 + MRC (hip) <3.
- Level 3: MRC (shoulder) ≥ 3 + MRC (hip) ≥ 3 + sitting at the edge of the bed without assistance (minimum 15 minutes).
- Level 4: standing without assistance and walking with or without assistance. Details of the early mobilization protocol based on Morris et al (1):

Level 1: Subjects with MRC <3 in the shoulder and hip admitted.

1. Sit on the bed with the headboard at a 60-80° angle (according to the subject’s tolerance).
2. Two sets of 10 repetitions of passive, active-assisted, or active movements (depending on the subject’s capacity) for the following joints:

Shoulder (abduction-adduction).

Elbow (flexion-extension).

Wrist (flexion-extension).

Hip (flexion-extension).

Knee (flexion-extension).

Ankle (dorsiflexion-extension).

Repetitions were performed with a full range of motion, respecting the subject’s optimal level, bilaterally.

Level 2: Subjects included with MRC ≥ 3 in the shoulder and <3 in the hip.

1. Two sets of 10 repetitions of active or assisted movements (depending on the subject’s capacity) for the following joints:

Shoulder (abduction-adduction).

Elbow (flexion-extension).

Wrist (flexion-extension).

Hip (flexion-extension).

Knee (flexion-extension).

Ankle (dorsiflexion-extension).

Repetitions were performed with a full range of motion, respecting the subject’s optimal level, bilaterally.

Level 3: Subjects with MRC (shoulder and hip) ≥ 3, who could tolerate sitting at the edge of the bed for at least 15 minutes without assistance.

1. We performed 2 sets of 10 repetitions of active movements while sitting at the edge of the bed for the following joints:

Shoulder (abduction-adduction).

Elbow (flexion-extension).

Wrist (flexion-extension).

Hip (flexion-extension).

Knee (flexion-extension).

Ankle (plantar flexion-dorsiflexion).

2. Assisted standing (according to the subject’s tolerance).

Level 4: Patients who were able to march in place were included.

1. Standing was performed without assistance and walking with or without assistance.

#### ETHICAL RESPONSIBILITIES

Protection of individuals and animals. The authors declare that no experiments have been conducted on humans or animals for this research.

Confidentiality of data. The authors declare that they have followed their workplace protocols regarding the publication of patient data.

Right to privacy and informed consent. The authors have obtained informed consent from the patients and/or subjects mentioned in the article. This document is in the possession of the corresponding author.

## References

1. Deem S. Intensive-Care-Unit-Acquired Muscle Weakness. Respir Care. 2006;51(9):1042–53.

2. Stevens RD, Marshall SA, Cornblath DR, Hoke A, Needham DM, De Jonghe B, et al. A framework for diagnosing and classifying intensive care unit-acquired weakness. Crit Care Med. 2009;37(10S):S299–308.

3. Schweickert WD, Hall J. ICU-Acquired Weakness. Chest. 2007;131(5):1541–9.

4. Lipshutz AKM, Gropper MA. Acquired Neuromuscular Weakness and Early Mobilization in the Intensive Care Unit. Anesthesiology. 2013;118(1):202–15.

5. Mendez-tellez PA, Needham DM. Early Physical Rehabilitation in the ICU and Ventilator Liberation. Respir Care. 2012;57(10):1663–9.

6. Garnacho-Montero J, Madrazo-Osuna J, García-Garmendia JL, Ortiz-Leyba C, Jiménez-Jiménez F, Barrero-Almodóvar A, et al. Critical illness polyneuropathy: risk factors and clinical consequences. A cohort study in septic patients. Intensive Care Med. 2001;27(8):1288–96.

7. van den Berghe G, Wouters P, Weekers F, Verwaest C, Bruyninckx F, Schetz M, et al. Intensive insulin therapy in critically ill patients. N Engl J Med. 2001;345(19):1359–67.

8. Hermans G, Wilmer A, Meersseman W, Milants I, Wouters PJ, Bobbaers H, et al. Impact of intensive insulin therapy on neuromuscular complications and ventilator dependency in the medical intensive care unit. Am J Respir Crit Care Med. 2007;175(5):480–9.

9. Bates B. The nervous system: a guide to physical examination and history taking. 5th ed. Philadelphia: J. B. Lippencott Company; 1991. 500–560 p.

10. Hough CL, Lieu BK, Caldwell ES. Manual muscle strength testing of critically ill patients: feasibility and interobserver agreement. Crit care. 2011;15(1):R43.

11. Hermans G, Clerckx B, Vanhullebusch T, Segers J, Vanpee G, Robbeets C, et al. Interobserver agreement of Medical Research Council sum-score and handgrip strength in the intensive care unit. Muscle Nerve. 2012;45(1):18–25.

12. Connolly BA, Jones GD, Curtis AA, Murphy PB, Douiri A, Hopkinson NS, et al. Clinical predictive value of manual muscle strength testing during critical illness: an observational cohort study. Crit Care. 2013 Oct;17(5):R229.

13. Fan E, Dowdy DW, Colantuoni E, Mendez-tellez PA, Sevransky JE, Shanholtz C, et al. Physical complications in acute lung injury survivors: A two-year longitudinal prospective study. Crit Care Med. 2014;42(4):849–59.

14. Hermans G, Van Mechelen H, Bruyninckx F, Vanhullebusch T, Clerckx B, Meersseman P, et al. Predictive value for weakness and 1-year mortality of screening electrophysiology tests in the ICU. Intensive Care Med. 2015;41(12):2138–48.

15. Gosselink R, Clerckx B, Robbeets C, Vanhullebusch T, Vanpee G, Segers J. Physiotherapy in the Intensive Care Unit. Netherlands J Crit Care. 2011;15(2):66–75.

16. Morris PE, Goad A, Thompson C, Taylor K, Harry B, Passmore L, et al. Early intensive care unit mobility therapy in the treatment of acute respiratory failure. Crit Care Med. 2008;36(8):2238–43.

17. Nordon-craft A, Schenkman M, Ridgeway K. Physical Therapy Management and Patient Outcomes following ICU-Acquired Weakness: A Case Series. J Neurol Phys Ther. 2012;35(3):133–40.

18. Rodriguez PO, Setten M, Maskin LP, Bonelli I, Romero S, Attie S, et al. Muscle weakness in septic patients requiring mechanical ventilation: Protective effect of transcutaneous neuromuscular electrical stimulation. J Crit Care. 2011;27(3):319.e1– 319.e8.

19. Karatzanos E, Gerovasili V, Zervakis D, Tripodaki E, Apostolou K, Vasileiadis I, et al. Electrical Muscle Stimulation: An Effective Form of Exercise and Early Mobilization to Preserve Muscle Strength in Critically Ill Patients. Crit Care Res Pract. 2012;2012:8.

20. Jones S, Wdc M, Gao W, Ij H, Wilcock A, Maddocks M. Neuromuscular electrical stimulation for muscle weakness in adults with advanced disease. Cochrane database Syst Rev. 2016;(10):Art. No.: CD009419.

21. Connolly B, Salisbury L, Neill OB, Geneen L, Douiri A, Mpw G, et al. Exercise rehabilitation following intensive care unit discharge for recovery from critical illness. Cochrane database Syst Rev. 2015;(6):Art.No.: CD008632.

22. Routsi C, Gerovasili V, Vasileiadis I, Karatzanos E, Pitsolis T, Tripodaki E, et al. Electrical muscle stimulation prevents critical illness polyneuromyopathy: a randomized parallel intervention trial. Crit Care. 2010;14(2):R74.

23. Schweickert WD, Pohlman MC, Pohlman AS, Nigos C, Pawlik AJ, Esbrook CL, et al. Early physical and occupational therapy in mechanically ventilated, critically ill patients: a randomised controlled trial. Lancet. 2009;373(9678):1874–82.

24. Bailey P, Thomsen GE, Spuhler VJ, Blair R, Jewkes J, Bezdjian L, et al. Early activity is feasible and safe in respiratory failure patients*. 2007;35(1):139–45.

25. Moss M, Nordon-Craft A, Malone D, Van Pelt D, Frankel SK, Warner ML, et al. A Randomized Trial of an Intensive Physical Therapy Program for Patients with Acute Respiratory Failure. Am J Respir Crit Care Med. 2015 Dec 10;193(10):1101–10.

26. Marra A, Ely EW, Pandharipande PP, Patel MB. The ABCDEF Bundle in Critical Care. Crit Care Clin. 2017;33(c):225–43.

